# The impact of isolation measures due to COVID-19 on energy intake and physical activity levels in Australian university students

**DOI:** 10.1101/2020.05.10.20076414

**Authors:** Linda A. Gallo, Tania F. Gallo, Sophia L. Young, Karen M. Moritz, Lisa K. Akison

## Abstract

**Background:** The pandemic inflicted by coronavirus disease 2019 (COVID-19) resulted in physical isolation measures in many parts of the world. In Australia, nationwide restrictions included staying at home, unless seeking medical care, providing care, purchasing food, undertaking exercise, or attending work in an essential service. All undergraduate university classes transitioned to online, mostly home-based learning. This disruption to daily life may have consequences for eating and physical activity patterns.

**Methods:** In this observational study, we examined the effect of isolation measures, during the early phase of the COVID-19 pandemic in Australia (March/April), on diet (24-hour diet recall, ASA-24) and physical activity (Active Australia Survey) patterns among third-year biomedical students in Brisbane, Australia. Findings were compared to students enrolled in the same course in the previous two years.

**Results:** In females, energy intake was ~20% greater in 2020 compared with 2018 and 2019, and the frequency of snacking and energy density of consumed snacks were also increased. In males, there was no difference in energy intake or snacking behaviour. Physical activity was impacted for both sexes, whereby fewer students undertook any walking activity and, of those that did, time spent doing so was less compared with 2018 and 2019. The proportion of students reporting any vigorous activity was not different for males or females but, among males who participated in this level of activity, the duration was less in 2020 compared with previous years. The proportion of male and female students achieving ‘sufficient’ levels of activity, defined by at least 150 mins over at least 5 sessions, was ~30% less in 2020. Indeed, the majority of students reported as having undertaken less physical activity than usual.

**Conclusions:** Increased energy intake for females and reduced physical activity for males and females demonstrate impacts of isolation measures that may have deleterious consequences for physical and mental wellbeing, with the potential to affect long-term nutrition and activity patterns.

## Introduction

Coronavirus disease 2019 (COVID-19), caused by SARS-CoV-2, was declared a pandemic by the World Health Organization on March 11^th^ 2020. In the majority of cases, it is an acute respiratory illness including fever, cough, and a sore throat but, in moderate to severe cases, the disease progresses to breathing difficulties, respiratory distress and extra-respiratory symptoms including heart and kidney injury and, in some cases, death (1). As of May 10^th^ 2020, the virus has already infected ~4 million people and resulted in more than 270,000 deaths globally (2).

Human-to-human transmission occurs primarily via respiratory droplets generated through coughing, sneezing, and talking; and other contagion sources include contaminated biologicals and surfaces (3, 4). Intense research efforts are in place for the identification of effective therapies and vaccines but, in the meantime, to contain spread and prevent overburdening our healthcare systems, the most effective strategy is contact tracing and physical isolation measures.

The Australian Government, including individual States and Territories, announced gradual ‘lockdown’ measures in response to the growing number of cases that could not be adequately traced. Community transmission become of major concern and, as at March 30^th^ 2020, all but essential services were shut down, and people only left their homes for work (in an essential service), or to purchase food, receive or provide medical care, or exercise. Whilst the majority of primary and secondary schools remained opened for the children of essential workers, universities transitioned all undergraduate learning online by March 23^rd^ 2020.

Whilst effective for containing outbreaks, disrupted habits involving strict isolation measures can adversely affect both physical and mental health, with potentially exacerbated effects among young adults who rely upon positive peer interactions for their general wellbeing (5, 6). Likely consequences of home-isolation are changes to eating and physical activity behaviours. It is reasonable to hypothesise that more time spent at home promotes hypercaloric diets, including larger meal sizes and increased snack frequency and size. Alternatively, fewer opportunities to travel outside the home may encourage more structured meal patterns and less take-away foods that are typically energy-dense. For physical activity, the lockdown would be anticipated to reduce both frequency and duration, not only for typically active persons who are no longer able to access gyms and health clubs, but for those who achieve sufficient levels of activity incidentally, through walking or cycling to work or study. This is particularly pertinent to university students attending a major campus who typically walk between classes numerous times a day. To this end, the impact of COVID-19-induced isolation measures on diet and physical activity patterns in Australian undergraduate students was measured one week after the transition to online learning and compared with data obtained in the previous two years.

## Methods

### Study design and participants

This observational study was approved by The University of Queensland Human Research Ethics Committee (Project Approval: 2016-02-066-PRE-3) and conducted in accordance with the National Statement on Ethical Conduct in Human Research (Australia).

Participants were recruited from third-year biomedical practical classes from The University of Queensland (Brisbane, Australia) in 2018, 2019 and 2020. In 2018 and 2019, students physically attended the practical classes on campus. In 2020, all undergraduate classes transitioned online by March 23^rd^. For each year, students who provided written informed consent were given a unique code (and password for the online diet questionnaire). Inclusion criteria was 19-27 years of age (214 males and 295 females).

### Diet and physical activity questionnaires

The Automated Self-Administered Dietary Assessment Tool (ASA24-Australia-2016) was used to guide participants through a 24-hour recall for the previous day (7). Participants were asked to recall all foods, drinks, and supplements consumed from midnight-to-midnight. Participants selected an eating occasion from a pre-determined list (e.g., ‘breakfast’ or ‘snack’) and reported all foods and beverages consumed at that time. Foods and beverages were entered by typing in specific search terms and selecting items from a returned list. Details of food types, preparation methods, portion sizes, additions, eating location, and food source were then queried by the system. Participants were prompted to recall frequently omitted and forgotten foods, and to complete a final review of all items consumed. The 24-hour energy intake included in this analysis includes all foods, drinks, and supplements. The main meals data include anything reported as ‘breakfast/brunch’, ‘lunch’ and ‘dinner’. Snack data include anything consumed during a ‘snack’, ‘drink’, and ‘supper’ occasion, which was before, after, or between main meals. Plain water or zero calorie drinks on their own were not considered a snack occasion. The distributions presented for eating location includes all occasions reported for all participants. For all meals eaten at home, the food source for the majority of ingredients reported for a given eating occasion is shown. For each year, the intake date used for the recall was between: March 18-20 2018, March 24-26 2019, and March 28-April 2 2020.

The Active Australia Survey was used to estimate leisure-time physical activity (8). Participants self-completed the survey following instruction. Sufficient activity was defined as at least 150 minutes of activity over at least five sessions per week. Insufficient activity was defined as undertaking some activity but not enough in total time or number of sessions to be considered a health benefit. Sedentary was defined as no activity at all. Total time was calculated by adding time spent in walking (continuously for at least 10 minutes), moderate activity and vigorous activity (weighted by two). To distinguish vigorous *versus* moderate activity, exercise intensity was estimated for each activity based on a metabolic equivalent (MET) score of 3-6 for moderate or >6 for vigorous, where 1 MET was defined as the resting metabolic rate, equivalent to oxygen uptake of 3.5 mL/kg/hour (9). A score for total sessions was calculated by adding the number of sessions of walking (continuously for at least 10 minutes), moderate activity, and vigorous activity. The average time spent walking (continuously for at least 10 minutes) or engaging in vigorous activity, for those who reported participation in that activity, was also reported, according to the Survey guide (8). For all calculations, any reported gardening or yard work was not included, as per survey guidelines.

### Statistical analyses

All analyses were performed using GraphPad Prism. Exploratory analyses were conducted, separately for males and females, examining the distribution and summary statistics. Where data did not follow a normal distribution, determined using the Shapiro-Wilk test, non-parametric tests were used, and data reported as median and either inter-quartile range (IQR) and/or range. Categorical variables were reported with frequencies and proportions. Distributions for participant ethnicity and dietary recalls that fell on a weekend were compared between years using the chi-square *(X^2^)* test. Due to non-normal data, a Kruskal-Wallis test was used to determine differences in age, between class years, followed by Dunn’s multiple comparisons test.

Due to non-normal data, a Kruskal-Wallis test was used to determine differences in total 24-hour energy intake (kJ), 24-hour energy intake and energy density attributed to main meals or snacks, and the number of snacking occasions between class years, followed by Dunn’s multiple comparisons test, where appropriate. As there were no statistical differences between 2018 and 2019, these years were combined and compared with class year 2020 for the same dietary parameters using a Mann-Whitney test. The proportion of participants reporting any alcohol consumption was compared between the years using the chi-square test. The location of eating occasions was compared between class years, using the chi-square test. As differences were found for both sexes, separate chi-square tests compared the ‘home’ location to all other locations combined. Given no statistically significant differences were found between class year 2018 and 2019, these years were combined and compared with 2020 using a further chi-square test. For all eating occasions at ‘home’, the source of food was compared between years, using the chi-square test.

To investigate differences in physical activity participation between class years, the proportion of respondents reporting any amount of walking and/or vigorous activity was examined. In order to meet the assumptions of the chi-square test, that all values are >1 and that no more than 20% of values are <5, a systematic approach was taken. Class year 2018 was compared to 2019 using the chi-square test and, given no statistically significant difference was found, these class years were combined and compared to 2020 using a separate chi-square test. Time spent in walking and vigorous activity was compared between class years using a Kruskal-Wallis test, followed by Dunn’s multiple comparisons test, where appropriate. Influence of class year on proportion of participants achieving ‘sufficient’ activity was also examined. Class year 2018 was compared to 2019 using a chi-square test and, given no statistically significant difference was found, these class years were combined. Sedentary and insufficient activity were also combined, resulting in a separate chi-square test comparing 2018/19 to 2020 for sedentary/insufficient *versus* sufficient levels of physical activity.

## Results

### Study participants

Table **1** shows the sample size for each outcome variable in males and females, and the proportion of dietary recalls that fell on a weekend relative to a weekday. This differed for both sexes between 2018 and 2019 due to differences in practical class scheduling (69% of students in 2018 and 36% in 2019 were scheduled on a Monday and therefore reported on meals and drinks consumed on Sunday; males: *X*^2^ (1) = 9.042, *p* < 0.01; females: *X^2^* (1) = 31.72, *p* < 0.0001). In 2020, students did not attend campus and had the option to complete the virtual practical class on any day within the specified week. In 2020, 31% of students reported weekend intakes which was significantly less compared with students in 2018 (males: *X*^2^ (1) = 23.52*, p* < 0.0001; females: *X^2^* (1) = 21.00*,p* < 0.0001) but not compared with 2019 (males: *X*^2^ (1) = 3.723, *p* = 0.054; females: *X^2^* (1) = 0.7808, *p =* 0.377). There were no differences in energy intake between weekday and weekend recalls for each year among males and females (data not shown; Student’s unpaired t-test or Mann-Whitney test, where appropriate).

**Table 1.** Participants included in the study

|  | Year 2018 | Year 2019 | Year 2020 |
| --- | --- | --- | --- |
| **Males** |  |  |  |
| Entered study (N) | 75 | 80 | 70 |
| >27 years of age or unknown (N) | 1 | 2 | 4 |
| No diet and physical activity data (N) | 3 | 1 | 0 |
| **Total included in study (N)** | **71** | **77** | **66** |
| **Included for diet [N, (%)]** | **65 (91.5)** | **63 (81.8)** | **64 (97.0)** |
| Missing diet data (N) | 6 | 14 | 2 |
| Weekend:weekday diet recall [N, (% weekend)] | 45:20 (69.2) | 27:36 (42.9)** | 17:47 (26.6)**** |
| **Included for physical activity [N, (%)]** | **61 (85.9)** | **73 (94.8)** | **66 (100)** |
| Missing physical activity data (N) | 10 | 4 | 0 |
| **Females** |  |  |  |
| Entered study (N) | 105 | 112 | 89 |
| >27 years of age or unknown (N) | 2 | 2 | 5 |
| No diet and physical activity data (N) | 0 | 2 | 0 |
| **Total included in study (N)** | **103** | **108** | **84** |
| **Included for diet [N, (%)]** | **101 (98.1)** | **96 (88.9)** | **82 (97.6)** |
| Missing diet data (N) | 2 | 12 | 2 |
| Weekend:weekday diet recall [N, (% weekend)] | 70:31 (69.3) | 28:68 (29.2)**** | 29:53 (35.4)**** |
| **Included for physical activity [N, (%)]** | **97 (94.2)** | **104 (96.3)** | **83 (98.8)** |
| Missing physical activity data (N) | 6 | 4 | 1 |

\*\* $p < 0.01$ vs. 2018, \*\*\*\* $p < 0.0001$ vs. 2018.

Table **2** summarises the demographic characteristics by class year. A significant age difference was found between class year for both males (H(2) = 27.37, *p* < 0.0001) and females (H(2) = 22.79, *p* < 0.0001). For males, median age was significantly higher in year 2020 compared with 2019 *(p* < 0.01) and 2018 *(p* < 0.0001). Similarly, for females, median age was significantly higher in class year 2020 compared with 2019 *(p* < 0.0001) and 2018 *(p* < 0.01). There was no difference in age between 2018 and 2019 for males or females. A chi-square test revealed no significant differences in ethnicity proportions between collection years for both males (X^2^ (8) = 11.73, *p* = 0.164) and females (X^2^ (8) = 7.271*,p* = 0.508).

**Table 2.** Participant demographics

|  | Year 2018 | Year 2019 | Year 2020 |
| --- | --- | --- | --- |
| **Males** | N=71 | N=77 | N=66 |
| Age [median (range), years] | 19 (19-25) | 20 (19-25) | 20 (19-27)*^ |
| Ethnicity [N (%)] |  |  |  |
| Asian | 23 (32.4%) | 18 (23.4%) | 25 (37.9%) |
| Asian sub-continental | 4 (5.6%) | 7 (9.1%) | 6 (9.1%) |
| Caucasian | 39 (54.9%) | 42 (54.5%) | 29 (43.9%) |
| Multi | 2 (2.8%) | 1 (1.3%) | 4 (6.1%) |
| Other/Not disclosed | 3/0 (4.2%) | 5/4 (11.7%) | 2/0 (3.0%) |
| **Females** | N=103 | N=108 | N=84 |
| Age [median (range), years] | 20 (19-26) | 20 (19-23) | 21 (19-26)*^ |
| Ethnicity [N (%)] |  |  |  |
| Asian | 29 (28.2%) | 28 (25.9%) | 27 (32.1%) |
| Asian sub-continental | 3 (2.9%) | 10 (9.3%) | 8 (9.5%) |
| Caucasian | 65 (63.1%) | 60 (55.6%) | 44 (52.4%) |
| Multi | 3 (2.9%) | 6 (5.6%) | 2 (2.4%) |
| Other/Not disclosed | 3/0 (2.9%) | 3/1 (3.7%) | 2/1 (3.6%) |

\* $p < 0.05$ vs. 2018, ^ $p < 0.05$ vs. 2019.

### Dietary intake from ASA 24-hour recall

For male participants, total 24-hour energy intake was not different between class years (H(2) = 0.992, *p* = 0.609; Figure **1A**). For females, a significant difference was found (H(2) = 7.60, *p* < 0.05), whereby total 24-hour energy intake during the COVID-19 pandemic in 2020 tended to be higher compared with the year 2019 (+13.2%; *p* = 0.067) and was significantly higher compared with the year 2018 (+24.3%; *p* < 0.05; Figure **1B**). As there was no significant difference between 2018 and 2019, these years were combined and compared with 2020. Among males, there remained no difference (Figure **1C**) and, among females, total 24-hour energy intake was 19.5% higher in 2020 compared with 2018/19 combined *(p* < 0.01; Figure **1D**).

**Figure 1.**
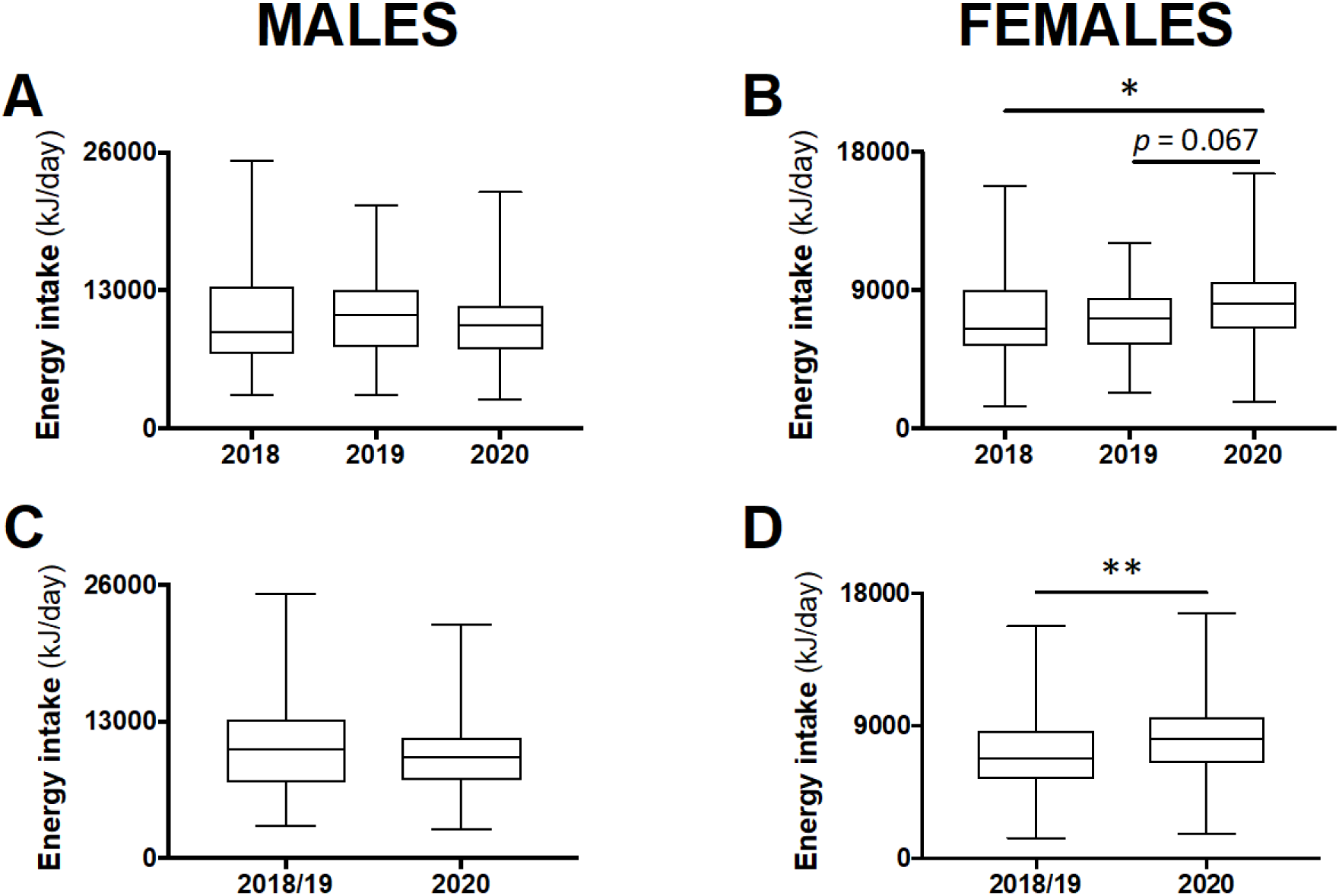
Total 24-hour energy intake in male and female students in class year 2020 (N=64 males, 82 females) compared with 2018 (N=65 males, 101 females) and 2019 (N=63 males, 96 females; A, B). No statistical differences were observed between 2018 and 2019 and therefore these years were combined (N=128 males, 197 females) and compared with 2020 (C, D). Data are presented as median ± IQR and range. * *p < 0.05* between 2020 and 2018 by Kruskal-Wallis test. ** *p < 0.01* between 2020 and 2018/19 by Student’s unpaired *t*-test.

Daily energy intake and energy density coming from main meals or from snacks were compared between the class years. There were no significant differences between 2018 and 2019, and thus these years were combined and compared with 2020. Daily energy intake from main meals was not different, but energy density tended to be less in male (−5.2%; *p* = 0.068), but not female, participants in 2020 compared with 2018/19 (Figure **2A-D**). Among males, there was no difference in the number of snack occasions, energy intake attributed to snacks, or the energy density of consumed snacks between 2020 and 2018/19 (Figure **2E, G, I**). However, in females, there was an increase to two snack occasions in 2020 compared with one in 2018/19 *(p* < 0.05; Figure **2F**). In addition, energy intake and energy density attributed to snacks were increased in 2020 compared with 2018/19 among female students (+50%; *p* = 083 and +62%; *p* < 0.05, respectively; Figure **2H, J**).

**Figure 2.**
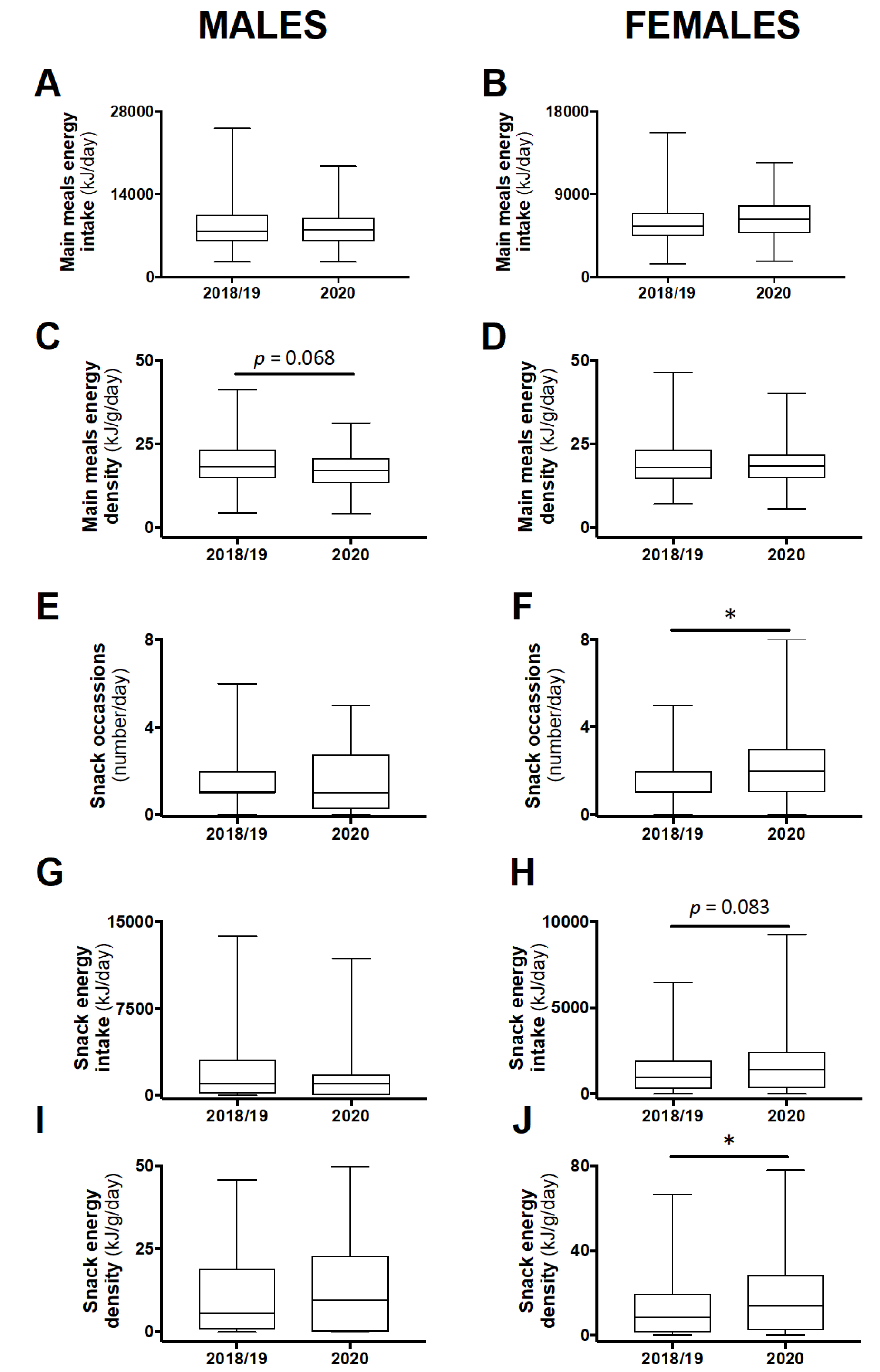
24-hour energy intake and energy density attributed to main meals (A-D) or snacks (E-H), and snacking frequency (I, J) in male and female students in class year 2020 (N=64 males, 82 females) compared with 2018/19 (N=128 males, 197 females). No statistical differences were observed between 2018 and 2019 and therefore these years were combined. Data are presented as median ± IQR and range. * *p < 0.05* between 2020 and 2018/19 by Student’s unpaired *t*-test.

The proportion of participants who consumed any alcohol between the class years was not different for males (19% average across all years, *X*^2^ (2) = 2.297, *p* = 0.317) or females (13% average across all years, *X*^2^ (2) = 3.425, *p* = 0.180; data not shown).

The distribution of eating location by class year was significantly different for both males (X^2^(8) = 71.27, *p* < 0.0001) and females (*X*^2^(8) = 86.53, *p* < 0.0001). Separate chi-square tests comparing the ‘home’ location to all other locations revealed a significant difference, with the vast majority of participants consuming food at home (males: 96.2% in 2020 and 73.8% in 2018/19 combined; *X*^2^(1) = 62.37,*p* < 0.0001; females: 95.8% in 2020 and 75.5% in 2018/19 combined; *X*^2^(1) = 78.33,*p* < 0.0001; Figure **3A-B**). For all foods consumed at home, the food source was not different between class years (males: *X*^2^(8) = 5.957, *p* = 0.652; females: *X*^2^(8) = 14.12, *p* = 0.079; Figure **3C-D**).

**Figure 3.**
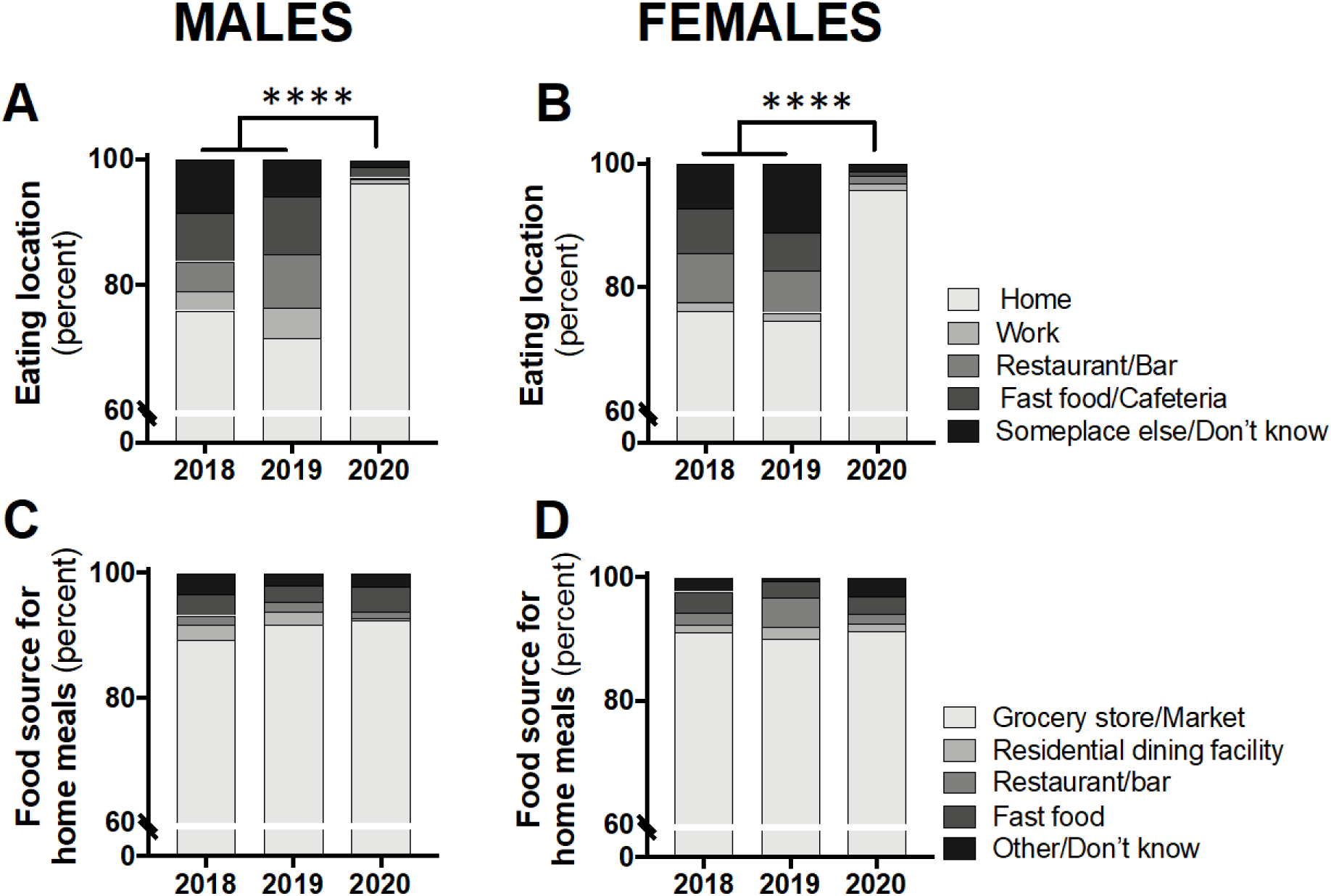
Eating location (A, B) and food source for all eating occasions at home (C, D) in male and female students in class year 2020 (N=66 males, 83 females) compared with 2018 (N=61 males, 97 females) and 2019 (N=73 males, 104 females). Data are presented as the proportion of students included in this analysis each year. No statistical differences were observed between 2018 and 2019 and therefore these years were combined for statistical comparisons with 2020. **** *p < 0.0001* between 2020 and 2018/19 for ‘home’ *vs*. all other locations by chi-square test.

### Physical activity levels from the Active Australia Survey

The proportion of participants reporting any amount of physical activity, for walking and vigorous, can be seen in Figure **4A-B**. In males, a chi-square analysis comparing year 2020 to 2018/19 combined revealed a significant reduction in walking participation *(X*^2^(1) = 3.969, *p* < 0.05) but no difference for vigorous activity (*X*^2^(1) = 1.287, *p* = 0.257; Figure **4A**). A similar result was seen for females with a significant reduction for walking participation in 2020 compared with 2018/19 *(X*^2^(1) = 6.299, *p* < 0.05) but no difference for vigorous activity *(X*^2^(1) = 1.35, *p* = 0.245; Figure **4B**).

**Figure 4.**
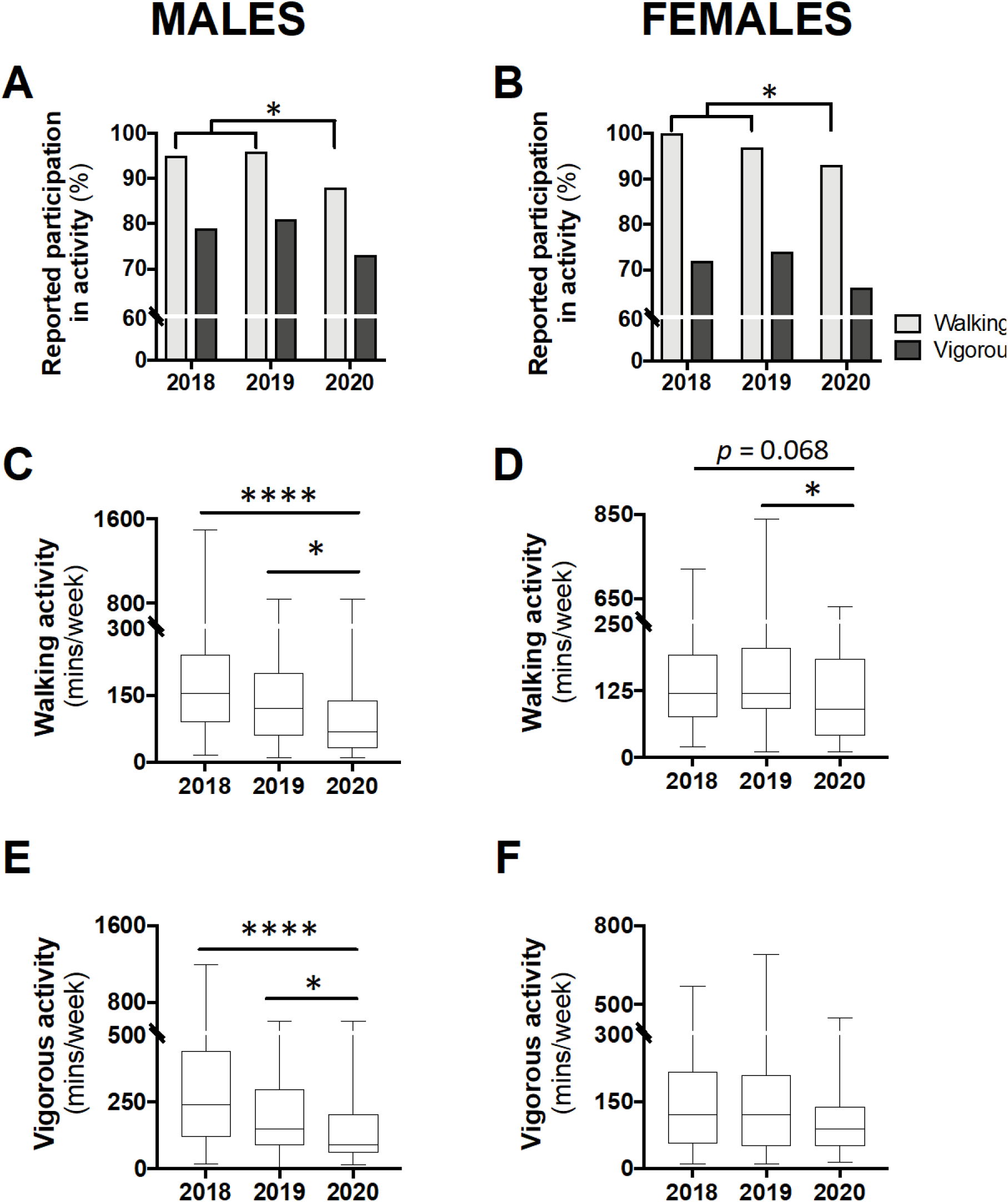
Levels of participation in physical activity (A, B) and time spent in walking (C, D) or vigorous activity (E, F) in class year 2020 (N=66 males, 83 females) compared with 2018 (N=61 males, 97 females) and 2019 (N=73 males, 104 females). Data are presented as the proportion of students included in this analysis each year (A, B), or median ± IQR and range (C-F). For A-B, no statistical differences were observed between 2018 and 2019 and therefore, to meet the assumptions of the chi-square test, these years were combined and compared with 2020; * *p < 0.05* between 2020 and 2018/19 by chi-square test. For C-E, * *p < 0.05* between 2020 and 2019 and **** *p < 0.0001* between 2020 and 2018 by Kruskal-Wallis test.

Among those who reported any walking, a Kruskal-Wallis test revealed a significant difference in the time spent in this activity for both males (H(2) = 19.11, *p* < 0.0001) and females (H(2) = 8.964, *p* < 0.05). Post-hoc analysis for male participants revealed that time spent walking was significantly less in year 2020 compared with 2019 (−52.5 min; *p* < 0.05) and 2018 (−87.5 min; *p* < 0.0001; Figure **4C**). Similarly, for females, time spent walking in 2020 was significantly less than 2019 (−30 min; *p* < 0.05) and non-significantly less than 2018 (−30 min; *p* = 0.068; Figure **4D**). Time spent walking between class years 2018 and 2019 were not different for either males or females. Time spent in vigorous activity was also found to be significantly different for males (H(2) = 19.63, *p* < 0.0001), with class year 2020 spending significantly less time in this activity compared with 2019 (−60 min; *p* < 0.05) and 2018 (−150 min; *p* < 0.0001; Figure **4E**). There was no difference in time spent in vigorous activity between males in year 2018 and 2019. Additionally, among females, no differences in time spent in vigorous activity were seen between the class years (Figure **4F**).

The proportion of participants achieving ‘sufficient’ activity, relative to insufficient and sedentary levels can be seen in Figure **5A-B**. A chi-square analysis comparing class year 2020 to 2018/19, revealed a significant difference for males (*X*^2^(1) = 13.36,*p* < 0.001; Figure **5A**) and females (*X*^2^(1) = 20.88, *p* < 0.0001; Figure **5B**), whereby fewer participants achieved ‘sufficient’ levels of activity in 2020. Figure **5C-D** demonstrates the proportion of participants from class year 2020 reporting typical *versus* atypical amounts of physical activity during the COVID-19 pandemic; the majority (56% for males and 61% for females) reported less than usual physical activity levels.

**Figure 5.**
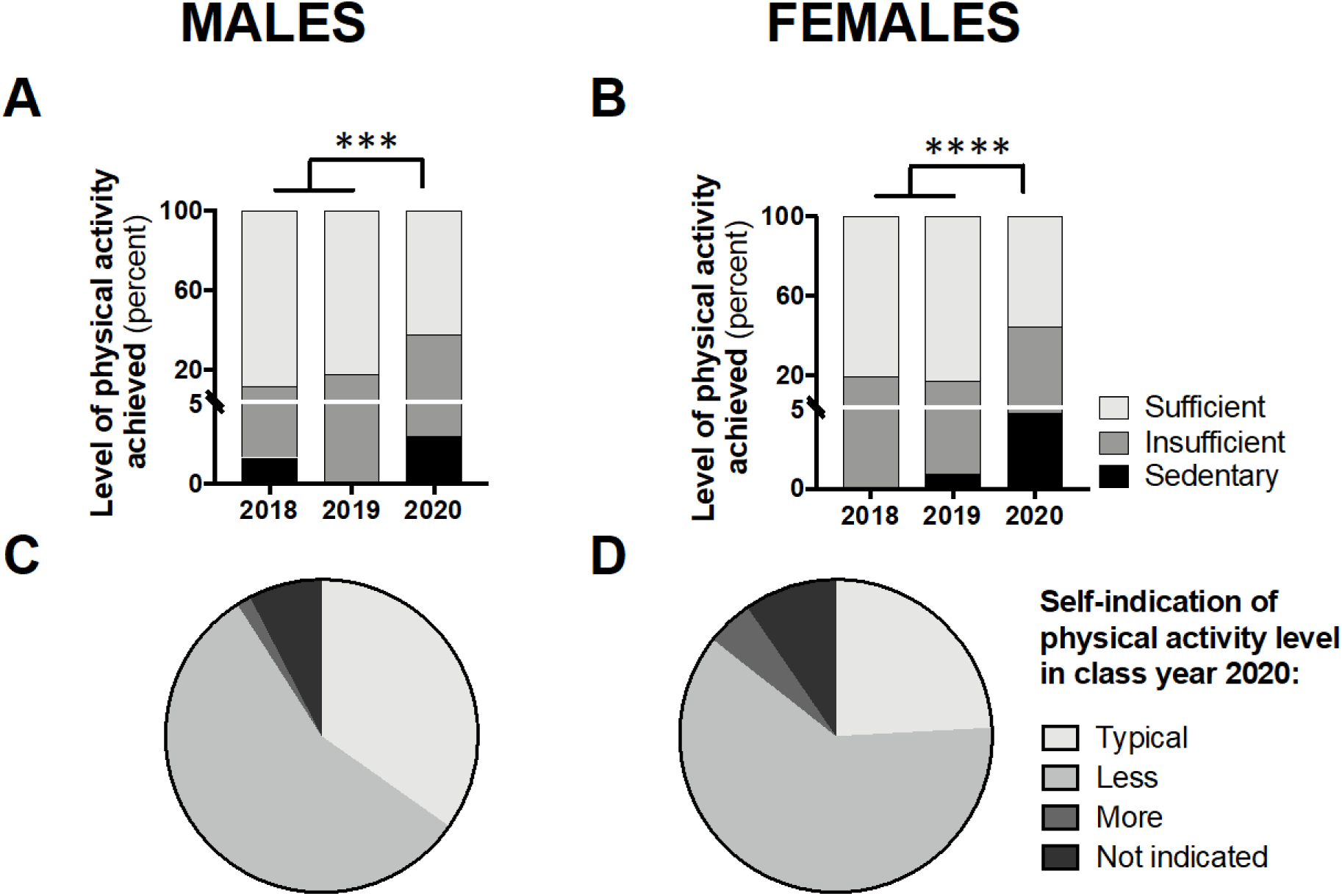
Students achieving ‘sufficient’ physical activity levels in class year 2020 (N=66 males, 83 females) compared with 2018 (N=61 males, 97 females) and 2019 (N=73 males, 104 females) (A, B) and students self-indicating ‘typical’ levels of physical activity in year 2020 (C, D). Data are presented as the proportion of students included in this analysis each year. For A-B, no statistical differences were observed between 2018 and 2019 and therefore, to meet the assumptions of the chi-square test, these years were combined and compared with 2020; *** *p < 0.001* and **** *p < 0.0001* between 2020 and 2018/19 for ‘sufficient’ *vs*. ‘sedentary/insufficient’ by chi-square test.

## Discussion

This is the first study to assess energy intake and physical activity levels in young adults during the COVID-19 pandemic. Our study was conducted in a cohort of Australian university undergraduate students within one week of government-imposed physical isolation measures and the transition from face-to-face to online classes. These data were compared with students enrolled in the same course over the preceding two years. The major findings are an increase in total energy intake, snacking frequency, and energy density of consumed snacks in female students, and a reduction in physical activity levels in both males and females.

Almost all (~96%) eating occasions during the pandemic were based at home compared with only three-quarters in 2018/19. This was expected, given the ban on dining-in at food venues that had remained open. Takeaway options remained available, but more than 90% of all foods consumed in the home were sourced from grocery stores and/or fresh markets, which was consistent with previous years. There was a small (~5%) reduction in the energy density of main meals consumed by male students in 2020 and, while the location of meal preparation was not examined in this study, this could reflect a reduction in foods prepared away from home, which are typically more energy dense compared with home-prepared foods (10). Total energy intake was unaffected, indicating a compensatory increase in main meal portion size. Compared with the difference in males, the maintenance of main meal energy density in females may reflect their higher consumption of home-prepared meals in general, pre-pandemic (11). Our data suggest, however, that more time spent at home promoted a hypercaloric diet in female students, which was attributed to increased frequency and energy density of snacks. Foods consumed away from home, including self-prepared lunches and snacks, are typically pre-portioned to a limited size. Thus, increased consumption at home may result from increased food visibility and opportunities to snack, possibly exaggerated by ‘panic buying’ and stockpiled food that coincided with national lockdown measures (12, 13).

Increased snacking in female, but not male, students while in home-isolation may reflect differences in social influence and perceived norms (14). When in the presence of mixed-sex peers, which is likely in the university setting, females have been shown to eat less than when in same-sex groups (15). Eating behaviour is also known to be impacted by stress and anxiety and, indeed, there has been a marked increase in telephone calls to youth mental health organisations during the pandemic, the majority from young women aged 19-25 (16). Females appear to be more likely to ‘stress-eat’ and consume hyperpalatable ‘comfort’ foods (17-19), which, in line with the current study, are typically energy dense. The addictive properties of energy-dense comfort foods can lead to long-lasting changes to eating behaviours (20).

As our reference control group, the median 24-hour energy intake in 2018/19 combined was 10,338 kJ for males and 6,776 kJ for females. This is slightly less than the latest data on energy intake from the Australian Health Survey, which included a broader representation of Australian residents (11,004 kJ in males and 7,863 kJ in females; means for 19-30 year age group) (21). Lower energy intake in biomedical science students compared with the general public may be due, in part, to greater health awareness and more balanced eating patterns. Alternatively, albeit somewhat related, is the established inverse relationship between education status and obesity in high-income nations (22). It should also be noted that the national nutrition data was attained in 2011-12, which may confound comparisons with the current data. Nevertheless, our data are consistent with a previous report in health science students in Spain (−22% and −8% compared with national survey data in males and females, respectively) (23), suggesting that university students consume less energy than age-matched national averages.

Isolation measures also saw a substantial reduction in physical activity levels in both male and female students, despite widespread recommendations to maintain physical activity during the pandemic (24, 25). Specifically, in 2020, fewer students reported any amount of walking (males 88%; females 93%) compared to 2018/19 (males 96%; 99% females), and of those that did, less time was spent doing so. It is reasonable to attribute this finding to the loss of incidental walking through commute, daily activities, or as part of one’s vocation, including walking between classes on campus. Those choosing to participate in vigorous activity, which was not different between the class years, are perhaps more ingrained in their prioritisation of physical activity (26) and found ways to continue doing so during lockdown. Notably, however, of those that did participate in vigorous activity, the time spent doing so in 2020 was less for males compared with previous years. With males more likely to participate in vigorous activity, as we and others have shown (27), and be motivated by mastery and competition (28), the closure of recreational sport and community gyms was likely to impact their activity levels.

The positive health benefits of physical activity are well established, with unequivocal evidence linking physical inactivity to non-communicable diseases (29). Many government bodies have established physical activity guidelines, not only as a prevention strategy for chronic diseases, but for psychological benefits (30-33). The most recent Australian Guidelines recommend adults between 18-64 years achieve at least 150 minutes of moderate intensity activity, on most days of the week, to achieve health benefits (34). In our population of undergraduate biomedical students, over 80% of participants were deemed to be ‘sufficiently active’ in the control years, which well-surpasses the age-controlled Australian average of 53% (35). While university educated persons have been reported to be more physically active, and have a reduced risk of being overweight (36), the reductions in sufficient activity seen during the COVID-19 pandemic, to 62% and 55% for males and females, respectively, is less than favourable. Specific targets have been set for ‘sufficient activity’ to achieve substantial health benefits, but the Australian Guidelines also stipulate that “doing any physical activity is better than doing none” (34). This is supported by research showing a dose-response relationship between positive health effects and physical activity, with no obvious lower threshold for benefit and a continuous risk reduction (37). Therefore, in this study, the increase in sedentary behaviour and reduction in physical activity raises health concerns.

While several weeks, or even a few months, of physical inactivity is unlikely to result in an abrupt onset of metabolic disease, sudden exercise cessation can decrease insulin sensitivity, cause muscle and bone loss, and abolish many of the positive exercise-induced metabolic and cardiovascular adaptations (38, 39). A further consequence is, undoubtedly, the loss of psychological benefits associated with physical activity (40). This may be compounded by the lack of social support during isolation periods, which is also important to maintain activity (41). This impact of inactivity on psychological health is further linked to the, potential stress-associated, increase in energy intake among female students. Arguably, the greatest risk to mortality, however, is the lasting impact on behaviour, that any time in lockdown may trigger. This is particularly true for those who are extrinsically motivated to participate in activity *(i.e*., to achieve outcome-based goals such as physical appearance) *versus* those who are intrinsically motivated and genuinely enjoy taking part in physical activity (42). It is worth noting that a hasty return to pre-pandemic activity levels, upon easing of isolation measures, can raise the risk of injury, particularly with the return to vigorous activity (43).

While these findings are undesirable, it is possible that our data reflect short-term consequences to an abrupt change in daily habits, and that over several weeks, participants recalibrated their diet and physical activity behaviours upon a newly established routine. For example, females may have started to consider the implications of increased snacking and both males and females may have taken the time to establish ‘at-home’ workouts, gain access to equipment, and/or incorporate a walk into their day. Indeed, in this study, 56% of males and 61% of females felt they had undertaken ‘less than typical’ physical activity levels during the first week of isolation and the transition to online study, and such self-awareness may increase the likelihood of positive behaviour adjustments.

## Conclusions

The global outbreak of COVID-19 resulted in restrictive isolation measures in many parts of the world. This was aimed at limiting transmission and hence reducing the burden on our healthcare systems, *i.e*. ‘flattening the curve’. In our study, which was conducted during the early phase of physical isolation and transition to online learning, energy intake in female students was increased and physical activity levels in both males and females was reduced compared with students in the previous two years. Undesirable changes to diet and physical activity patterns have the potential to persist for some time, even after isolation measures are eased. These changes, particularly if sustained, can have deleterious consequences for both physical and mental wellbeing.

## Data Availability

Available upon request

## Abbreviations

COVID-19: Coronavirus disease 2019
MET: Metabolic equivalents
SD: Standard deviation

## Declarations

### Ethics approval and consent to participate

This study was approved by The University of Queensland Human Research Ethics Committee (Project Approval: 2016-02-066-PRE-3) and conducted in accordance with the National Statement on Ethical Conduct in Human Research (Australia).

Informed written consent was obtained from the study participants.

### Consent for publication

Not applicable.

### Availability of data and materials

Available upon request.

### Authors’ contributions

LAG, KMM, and LKA conceptualized the study. LAG, SLY, and LKA performed the data collection. LAG and TFG completed the statistical analysis. LAG, TFG, and SLY drafted the manuscript. All authors read, reviewed, and approved the final version of the manuscript.

## Acknowledgments

We thank the student volunteers from BIOM3020 Integrated Endocrinology (The University of Queensland) who took part in the study.

